# Construction and Evaluation of China Elderly Care Service Smart Supply Chain System

**DOI:** 10.1101/2023.06.07.23291093

**Authors:** You-Yu Dai, Guanlong Liu

## Abstract

With the continuous deepening of the population aging degree, the pressure on social pensions has gradually increased, and the problem of pensions has become a must to be solved. With the development of science and technology, people’s living standard is constantly improving. The demand of the elderly group for old-age service is not only limited to the basic daily life of individuals before; it is moving toward the need for services that make people happier and healthier, how to integrate modern science and technology with the old-age service industry to make it serve the old-age industry better. This research is about constructing and evaluating the intelligent supply chain of Chinese old-age service. Based on the research results of previous scholars, this paper divides the construction of the intelligent supply chain of China endowment service into four aspects: policy aspect, economic aspect, social aspect, and technical aspect; the four significant aspects are divided into the first-level indicators, and 16 second-level indicators are divided under the first-level indicators. The importance and satisfaction of each evaluation index were obtained by distributing questionnaires to the managers who study the supply chain and the employees who are related to the old-age service. After the reliability analysis, the IPA quadrant analysis chart of the evaluation index was constructed using importance-performance analysis. The index of constructing an intelligent supply chain system for China old-age service is given priority, the supply chain system of China old-age service is further improved, and the social security of China old-age service is enlarged.

## 1 Introduction

With the continuous development of the social economy and technology, various fields have achieved unprecedented improvements in China. However, with the continuous deepening of population aging, the pressure on social elderly care is also increasing year by year, and the issue of elderly care has become a problem that needs to be solved in China and even China. At the same time, with the improvement of people’s living standards, older adults are no longer meeting their basic needs for elderly care services but are developing towards higher levels of health, longevity, and meeting their spiritual and cultural needs. So our current priority is to effectively integrate modern information technology with the elderly care service industry, create an intelligent supply chain system for elderly care services, and apply it to elderly care services in China to effectively alleviate the aging problem in China.

“The Opinions of the Central Committee of the Communist Party of China and the State Council on Strengthening the Work on Aging in the New Era” pointed out that the implementation of the national strategy of active population aging is related to the overall development of the country, the well-being of billions of people, and has significant and far-reaching significance for the sustained and healthy development of Chinese economy and society during the 14th Five Year Plan period and beyond. We should respond to population aging with a more positive attitude, policies, and actions so that every older person can live a peaceful life, Meditation and comfort. [1] In terms of the current elderly care situation, older adults have a higher level of pursuit for realizing their own life value and cultural, entertainment, and spiritual aspects, not only limited to meeting basic living needs such as catering and medical care in the past.

According to the latest data from the 7th National Population Census, as of November 1, 2020, the total population in China was 1411.78 million, including 264.02 million people aged 60 and above, accounting for 18.70% of the total population. Compared with the 6th National Population Census data in 2010, the increase was 5.44 percentage points. The aging population is becoming increasingly severe, but traditional elderly care models can no longer meet the current needs of elderly care. Traditional elderly care services include home, institutional, and community care. However, with the high cost and low quality of elderly care services becoming increasingly prominent, the existing operational models of elderly care services can no longer meet the diverse needs of the aging population. Based on the continuous popularization of information digitization, utilizing Internet technologies such as big data, cloud platforms, and the Internet of Things to achieve service combinations has become an effective way to innovate smart elderly care service models. By using modern information technology to re-integrate and allocate social resources and reasonably allocate them to various projects in the smart elderly care service supply chain, we aim to meet the spiritual and material needs of the elderly population with low-cost and low resources, alleviate the elderly care burden on families and society, and improve the quality of social elderly care services.

The issue of elderly care is not only the primary concern and needs to be solved in China but also a problem that the entire society needs to solve nationwide. At present, there is relatively little research on innovative elderly care services. Through continuous innovation and in-depth research on the intelligent supply chain of elderly care services in China, we can effectively solve elderly care problems and promote the social elderly care security theory in China, which has particular significance. The elderly population is different from other groups and belongs to a relatively vulnerable group in society. Therefore, the intelligent supply chain system for elderly care services should be more targeted and specific. By surveying older adults and staff serving the elderly population in China, the supply chain system for elderly care services in China can be further improved, thereby increasing the social security for elderly care in China. The elderly population and staff serving older adults should provide objective feedback on various information and indicators to make more sweeping changes and innovations to the intelligent supply chain system of elderly care services based on the feedback content and improve the living standards of older persons.

Establishing a comprehensive China elderly care service supply chain system can effectively improve the elderly care service capacity in China. The intelligent elderly care service supply chain system can integrate elderly care service resources through intelligent information technology, analyze how innovative elderly care services can better explore and meet the service needs of older persons, and further improve the simplicity, professionalism, and precision of elderly care services. It has specific reference significance for the relevant elderly care departments in China. By understanding and investigating the elderly population and service personnel serving them, we propose reasonable suggestions for the current development status and challenges of elderly care services, effectively utilize and integrate social resources, maximize the limited social resources, promote the construction of intelligent elderly care services in China, and improve the level and capacity of social elderly care security.

This article takes the intelligent supply chain system of elderly care services in China as the research object. It uses the questionnaire survey method for empirical analysis to study the intelligent supply chain system of elderly care services in China, proposing optimization paths for the intelligent supply chain system of elderly care services in China. The innovation lies in studying the current situation of intelligent elderly care services in China, analyzing the advantages and disadvantages of the current innovative supply chain system for elderly care services in China, and summarizing research factors related to policy, economy, society, and technology. Deepening research on these four aspects is extremely important in the intelligent supply chain system for elderly care services in China. At the same time, this article conducts investigations and evidence collection in four different directions and ultimately concludes.

## 2 Literature Review

The issue of population aging is not only severe in China but also in foreign countries. Therefore, both domestically and internationally, efforts are being made to study the construction of an intelligent supply chain system for elderly care services. With the continuous development of technology and the economy, how can we utilize various modern high-tech technologies to build a complete elderly care service system, Domestic and foreign people are constantly exploring and innovating.

The research on thoughtful elderly care in China started relatively late. In terms of the literature on intelligent elderly care services in domestic literature, the start of thoughtful elderly care in China is relatively late, but it is increasing year by year, and this field is also improving year by year.

Chen (2014) [4] conducted a survey and research on hundreds of older people in society, dividing home-smart elderly care into five different dimensions: daily care, economic support, safety protection, rehabilitation and health care, and spiritual comfort. Then, he constructed a high-quality, high-efficiency, and low-cost smart elderly care service to meet the development needs of an aging society.

Li (2015) [5] believed that a new intelligent elderly care service system should be constructed suitable for the current social development in China, and the critical role played by traditional elderly care models cannot be ignored. He advocates optimizing the allocation of various social resources, encouraging older people to strengthen self-learning, improve their acceptance, and meet older people’s elderly care service needs.

Meng (2017) [6] conducted a study on the practicality of social networking sites browsed by older adults and found that the social networking sites frequently browsed by older people currently have shortcomings. Therefore, it is necessary to continuously transform and upgrade these social networking sites to make their service functions more healthy and informative, enabling older persons to receive effective guidance and assistance when browsing social networking sites. Making social networking sites for older people more functional and scientific and better meet older adults’ convenient use of social networking sites.

Zeng and Hou (2021) [7] found that Chinese policies on innovative elderly care services are immature. The incentive policies for elderly care services need to be strengthened, and the review and supervision policies for the intelligent elderly care service system need to be improved to ensure the sustainable and healthy development of innovative elderly care.

Chen (2020) [8] built a competent elderly care comprehensive service platform and designed a conceptual model of the intelligent elderly care supply chain to provide constructive suggestions for effectively integrating various elements such as elderly care service node enterprises, elderly care logistics, elderly care fund flow, elderly care information flow, etc., guiding and stimulating elderly care demand, monitoring and improving elderly care supply, and ensuring the realization of the value co-creation and value appreciation of elderly care services. On this basis, design five mechanisms: integration mechanism, guidance mechanism, coordination mechanism, cooperation mechanism, and incentive mechanism to ensure the healthy, stable, and sustainable operation of the intelligent elderly care service supply chain.

Dong (2020) [9] With the development of elderly care socialization and the increasing demand for elderly care services, how to provide affordable elderly care services has become a problem that the government and various fields of society must consider. Therefore, it is necessary to integrate resources from all parties reasonably, build an excellent elderly care service supply chain that meets the needs of older persons, meet their elderly care needs, improve their quality of life, and achieve the rational allocation of resources and the integrated development of the elderly care machine logistics industry to avoid resource waste. The British Life Trust was the first to propose the concept of “smart retirement.” G. Demiris (2008) [10] pointed out that smart home care is a service model that uses intelligent service equipment and information technology as carriers to systematically provide remote monitoring services for older people further to enhance their self-care ability and elderly care level.

Lemlouma (2013) [11] constructed a dependent intelligent elderly care system framework and then analyzed the particular service provision projects and times for each older adult based on scientific evaluation, providing different services according to their different situations.

Chan and Campo (2009) [12] believed that innovative elderly care services should integrate multiple social resources to build an Internet home care service system centered on older persons, which includes a unified collection of related technologies, services, and information integration.

Many scholars have conducted in-depth research on the supply chain of smart elderly care services. According to the construction of smart elderly care service supply chains by many scholars, they can be divided into four aspects: factors from policies, factors from the economy, factors from society, and factors from technology. Among them, the policy factors include Che and Zhou’s proposal of improving the incentive mechanism in 2022 [13], standardizing process management, and conforming to the evaluation and supervision criteria. Chen proposed in 2020 that enterprise qualifications should be complete. Economic factors include Ma, Yi, and Hu’s proposal to ensure service profits in 2020 [14]; Liu proposed in 2022 [15] that the quality of employees should be high; Dong proposed in 2020 that product quality is guaranteed. Social factors include Wang and Chen’s proposal in 2022 [16] that the base of elderly care entities is large; Shen proposed in 2017 that the current development situation should be rapid; Chen proposed in 2020 that there should be more social elderly care enterprises; Li proposed in 2015 that the target audience should have a higher level of acceptance. Technological factors include Zhu’s proposal in 2012 [18] to promote the popularization of the Internet of Things; Demiris (2008) [10] proposed the deepening of artificial intelligence; Song proposed in 2022 that cloud computing responds quickly; Zeng and Hou proposed in 2021 that the scale of extensive data collection is large; Meng proposed in 2017 [6] that mobile communication has broad coverage.

Based on the issues and current situation of the intelligent elderly care service supply chain proposed by other scholars, this article summarizes that the China Bright elderly care service supply chain is divided into four parts, as shown in Table 1.

**Table 1.** Establish an indicator system.

|  |  |  |
| --- | --- | --- |
| Factors from policies | Improvement of incentive mechanism | Che & Zhou (2022) [13] |
|  | Completeness of enterprise qualifications | Chen (2020) [8] |
|  | Process management should be standardized. | Che & Zhou (2022) [13] |
|  | The evaluation and supervision criteria should be consistent | Che & Zhou (2022) [13] |
| Factors from Economy | Guaranteed service profit | Ma, Yi, & Hu, (2020) [14] |
|  | The quality of employees should be high | Liu (2022) [15] |
|  | Product quality guaranteed | Dong (2020) [9] |
| Factors from society | Large base of elderly care entities | Wang & Chen (2022) [16] |
|  | The current development situation needs to be rapid | Shen (2017) [17] |
|  | More social elderly care enterprises | Chen (2020) [8] |
|  | The acceptance level of the target audience | Li (2015) [5] |
| Factors from technology | The popularization of the Internet of Things | Zhu (2012) [18] |
|  | Deepening of artificial intelligence | Demiris (2008) [10] |
|  | Cloud computing responds quickly. | Song (2022) [19] |
|  | The large scale of extensive data collection | Zeng & Hou (2021) [7] |
|  | Broad coverage of mobile communication | Meng (2017) [6] |

### Summary

Supply chain performance evaluation refers to the pre, during, and post-analysis and evaluation of the entire supply chain, including various links (especially the operational status of core enterprises and the functional relationships between each link), centered around the goals of the supply chain. Evaluating the performance of the supply chain is an evaluation of the operational performance of the entire supply chain, the cooperation between supply chain node enterprises, and the node enterprises on the supply chain. Supply chain performance evaluation indicators are performance evaluation indicators based on business processes. Supply chain performance evaluation is an important content of supply chain management, which is of great significance for measuring the achievement of supply chain goals and providing business decision support. [20] This article is based on the analysis and research of various indicators for building an intelligent supply chain system for elderly care services in China, evaluating the operational effectiveness of the entire intelligent supply chain system for elderly care services, and ultimately obtaining the construction and evaluation of the intelligent supply chain system for elderly care services in China.

Due to the late start of innovative elderly care services and economic and technological factors in China, the intelligent supply chain system for elderly care services is still in its infancy. In this regard, there is a slight lack of theory and practice on competent elderly care. However, due to the rapid development of the economy and technology, as well as the continuous emphasis of the government on intelligent elderly care services, many regions have conducted research on innovative elderly care services, which play a significant role in the development of intelligent supply chain systems for elderly care services in China. Compared to the mature, intelligent supply chain systems for elderly care services in foreign countries, we still have a particular gap. In addition, due to the differences in policies between China and foreign countries, it is not possible to directly apply foreign theories of innovative elderly care services. We can only rely on ourselves to explore and find a path that is suitable for Chinese national conditions.

After the above research shows shortcomings in developing innovative elderly care services domestically and internationally. Firstly, since the theory of intelligent elderly care development has only just emerged in the past decade, all the tests it has gone through have not yet been thoroughly tested, and future practice is needed to enrich the theory and meet current development needs continuously. Secondly, due to the limitations of elderly care services, most studies mainly focus on the technical level of innovative elderly care services, and there is little research to analyze the acceptance of intelligent elderly care by the elderly population.

## 3 Research Design

The research object of this article is to construct and evaluate the intelligent supply chain for elderly care services in China. The research aims to analyze the intelligent supply chain system for elderly care services in China based on its four major aspects of construction and propose reasonable suggestions to optimize the intelligent supply chain system for elderly care services in China and alleviate the burden of elderly care in China. This article mainly divides the intelligent supply chain system for elderly care services in China into four major parts, which are from four aspects: policy, economy, society, and technology. At the same time, the second level title is extended according to these four aspects. For example, aspects of the policy include (improvement of incentive mechanism, integrity of enterprise qualification, standardization of process management, and consistency of evaluation and supervision criteria). The Delphi method and IPA method are taken as the theoretical basis.

### 3.1 Delphi method method

The primary indicators in this article include four indicators: factors from policies, factors from the economy, factors from society, and factors from technology. Firstly, a preliminary survey questionnaire was designed and distributed to experts related to the research on the intelligent supply chain system for elderly care services in China. Their processing results were obtained. Based on their processing results, the 80% pass rate indicator was retained, and indicators below 80% were modified until the pass rate exceeded 80%.

The selection methods for consulting experts in this article mainly include (1) practitioners engaged in China elderly care services, (2) social research scholars engaged in researching the intelligent supply chain of China elderly care services, and (3) school teachers engaged in researching the supply chain. 12 experts were selected for the consultation questionnaire.

This article selects four primary indicators and sixteen secondary indicators. The primary indicators set in this article include four primary indicators: factors from policies, factors from the economy, factors from society, and factors from technology. Based on the four primary indicators, sixteen are differentiated, and the secondary indicators correspond to the primary ones. Firstly, a preliminary survey questionnaire was designed and distributed to experts related to the research on the smart supply chain system for elderly care services in China. Their processing results were obtained. Based on their processing results, the 80% pass rate indicator was retained, and indicators below 80% were modified until the pass rate exceeded 80%.

### 3.2 Questionnaire Design

The questionnaire design in this article has two dimensions, one is essential, and the other is expressiveness. Each dimension has 20 questions, totaling 40 questions. Likert’s 5-Point scale was used to assess the importance and performance of building an intelligent supply chain system for elderly care services in China.

### 3.3 Sampling design

A survey questionnaire is a way to help obtain raw data, mainly expressed through standardized data and designed questioning methods, to achieve the purpose of the survey. This questionnaire is designed to construct a smart supply chain system for elderly care services in China, considering multiple factors such as research background and survey subjects and combining them with one’s research objectives. The survey in this study adopts a nonrandom sampling method, which is a judgmental sampling in nonrandom sampling. A questionnaire survey is distributed to individuals with specific experience or knowledge. As the research focuses on the construction and evaluation of the smart supply chain system for elderly care services in China, in order to ensure the authenticity, reliability, and authority of the research results, the scope of the research object is determined to include relevant personnel studying the smart supply chain system for elderly care services in China and workers participating in elderly care services in China. Survey questionnaire through online survey platform “Questionnaire Star”(https://www.wjx.cn/vm/tErMJyw.aspx#)Source of data obtained.

After the expert group discussed the survey questionnaire, it was believed that the overall content of the questionnaire design could build an intelligent supply chain system for elderly care services in China, and the questionnaire was only distributed. The survey questionnaire was distributed on November 29, 2022, and 198 questionnaires were collected. All valid questionnaires were sorted and collected, and data entry and statistical analysis were conducted using the statistical software SPSS 24.0. The questionnaire was divided into two parts:

The first part is to investigate the problems related to each variable in the construction of the China elderly care service intelligent supply chain system and put forward the relevant questions about the importance and performance of each variable to obtain the importance and expressiveness of each variable. The second part of the content should mainly investigate the questionnaire participants’ information, such as their age distribution, occupation, and years of work related to elderly care services.

### 3.4 IPA Statistical Methods

Importance&Performance Analysis (IPA) is the analysis of customers’ perception of the importance and performance of a particular process in order to find ways to improve consumer satisfaction and loyalty [21]. The main purpose of the importance expressiveness analysis method is to provide the characteristics of the enterprise’s product or service quality, which can provide opportunities for continuous improvement. Therefore, customers must evaluate the importance and expressiveness of the product or service quality characteristics through questionnaires.

## 4 Research Results

### 4.1 Descriptive Statistical Analysis

This survey questionnaire was officially conducted, and 198 copies were distributed. The specific population statistics are shown in Table 2.

**Table 2.** Demographics

|  | Number of people | percentage (%) |
| --- | --- | --- |
| Male | 88 | 44.44 |
| Female | 110 | 55.56 |
| Aged 21-30-year-old | 70 | 35.35 |
| Aged 31-40-year-old | 54 | 27.27 |
| Aged 41-50-year-old | 24 | 12.12 |
| Aged 51-60-year-old | 26 | 13.13 |
| Aged more than 60<br>years old | 24 | 12.12 |
| College/Junior<br>College | 106 | 53.54 |
| Master's degree | 60 | 30.3 |
| Doctor's degree | 32 | 16.16 |
| School Teacher | 70 | 35.35 |
| Enterprise staff | 84 | 42.42 |
| government agent | 44 | 22.22 |
| manager | 60 | 30.3 |
| Front-line staff | 94 | 47.47 |
| Government staff | 44 | 22.22 |
| Less than one year | 62 | 31.31 |
| 1-5 years | 60 | 30.3 |
| More than five years | 76 | 38.38 |

### 4.2 Reliability and Validity Analysis

#### 4.2.1 Reliability test

In this pre-survey, the importance and expressiveness are divided into four levels, namely, from the policy aspect, from the economic aspect, from the social aspect, and the technical aspect. Reliability analysis mainly evaluates the reliability of the overall questionnaire. The results of this study show that in terms of importance, Cronbach’s α The value is 0.949, while Cronbach’s of expressiveness α The value is 0.987. α The value must be at least greater than 0.5, and α A value more excellent than 0.7 indicates high reliability. The research results show that each level meets high-reliability criteria α The value is more than 0.7, so the measurement reliability of the importance and expressiveness of this questionnaire is good. The test results are detailed in Table 3.

**Table 3.**
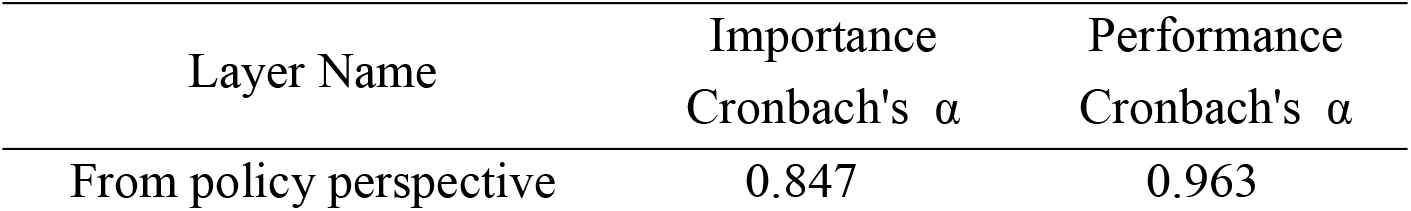
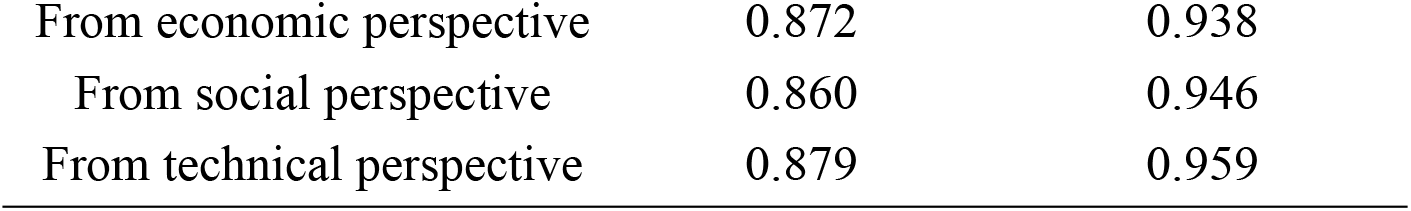
Cronbach’s alpha scale pre-survey

#### 4.2.2 Validity test

Expert validity: Determine the final indicator by combining relevant research from previous scholars and scoring experts based on questionnaires. Content validity: Based on an extensive review of previous literature, various factors are summarized to form a survey questionnaire. Experts and scholars then graded and tested whether the questionnaire design conforms to the research topic. Experts and scholars assisted in the clarity, face validity, and content validity of the questions in this scale, which helped to make the seal more accurate. Therefore, 12 participants conducted a content relevance assessment on constructing an intelligent supply chain system for elderly care services in China, evaluated the suitability between each cross-sectional area and the measurement questions, and checked whether the questions represented the module. The items were added or reduced through expert discussions, merged or differentiated, and evaluated and revised. The initial questionnaire was formed. Then distribute the questionnaire. The questionnaire results have good validity.

### 4.3 IPA Analysis of the Construction of China Elderly Care Service Smart Supply Chain System

This paper uses IPA (Importance&Performance Analysis) analysis to explore the expressiveness evaluation of each indicator for the construction of the China elderly care service intelligent supply chain system by taking the average of importance and performance as the dividing point, cutting the space into four quadrants on the X and Y axes, taking importance as the X axis and expressiveness as the Y axis.

In order to present the importance and expressiveness of each indicator in the construction of the China elderly care service intelligent supply chain system, this study revised the IPA analysis method studied by Chen Xu [22], so the overall average value of the importance and expressiveness of each indicator is taken as the standard to distinguish the four quadrants, so that the importance and performance of each indicator can be more intuitively compared. Therefore, the overall average value of 4.06 of importance is taken as the X coordinate, and the overall average value of 4.15 of expressiveness is taken as the Y coordinate as the central intersection point. A scatter diagram of four quadrants is made accordingly. See Table 4 and Figure 1 below for specific values.

**Table 4.**
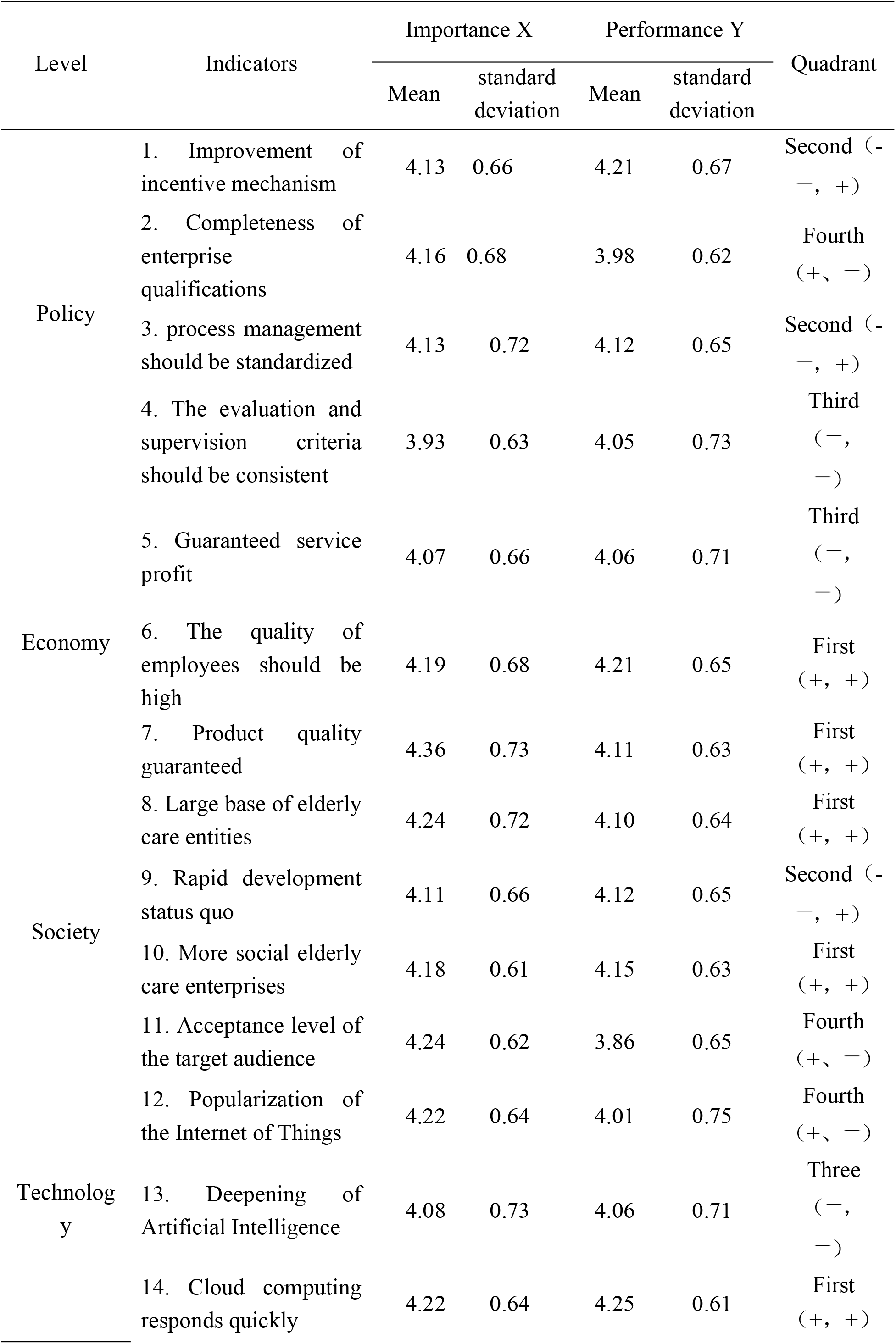
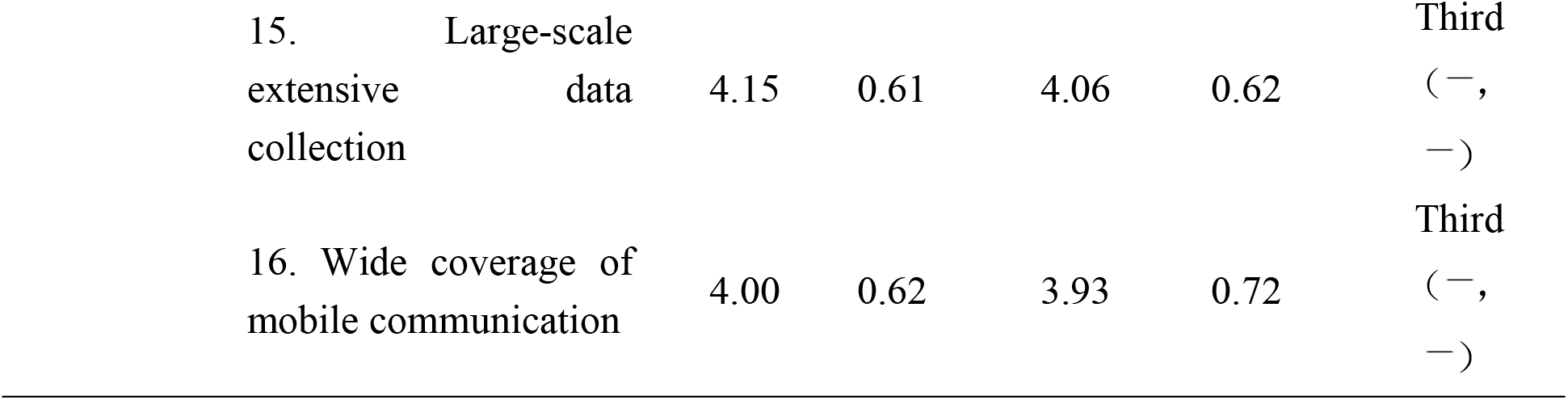
The coordinate values of each index

**Figure 1.**
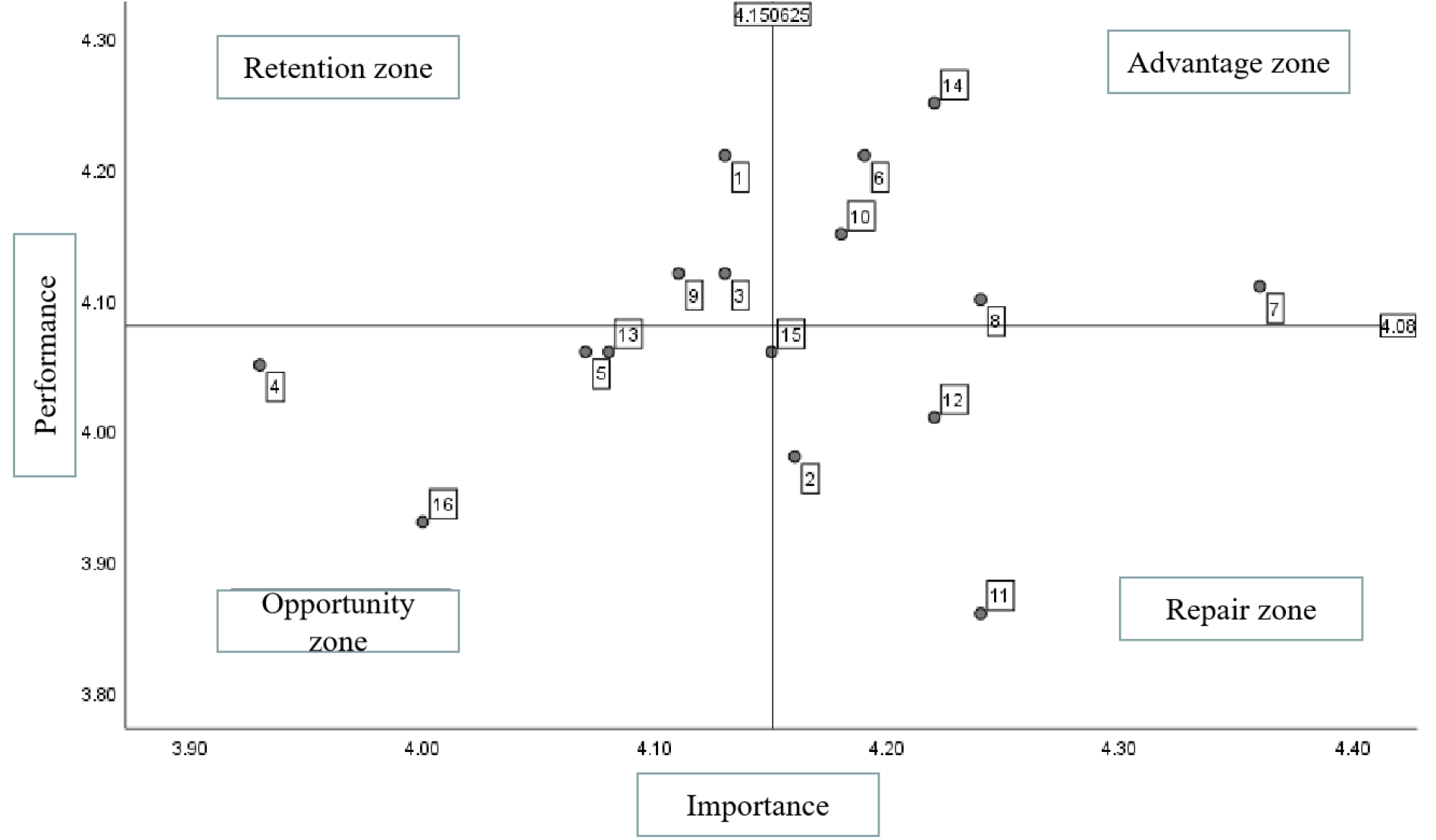
IPA analysis diagram

The IPA analysis chart can be divided into four quadrants. The first quadrant is the “advantage zone,” indicating that constructing an intelligent supply chain system for elderly care services in China is essential, and the results are satisfactory. Therefore, as long as the existing services are maintained, it is sufficient. The second quadrant is the “maintenance zone”, indicating that the construction of an intelligent supply chain system for elderly care services in China only needs to continue to maintain this element without investing excessive resources to avoid resource waste. The third quadrant is the “opportunity zone,” the improvement factors in this quadrant are a secondary focus, indicating that this element is not very important and does not need to be urgently addressed. The fourth quadrant is the “repair area,” indicating that relevant personnel believes that this aspect is essential in building an intelligent supply chain system for elderly care services in China, but there has been no good improvement, so priority must be given to improvement and strengthening.

According to the analysis of the secondary evaluation indicators, in terms of policy, the importance of improving incentive qualification and process management in the intelligent supply chain system of China elderly care service is not high. However, the expressiveness is relatively high, which indicates that the current situation of China elderly care is more suitable and only needs to continue to maintain and improved. The consistent importance and expressiveness of the criteria for evaluating and supervising innovative elderly care services are not high. This indicates that its proportion in the supply chain of Chinese innovative elderly care services is unimportant and can be postponed. The importance of improving the qualification of elderly care enterprises is relatively high, but the expressiveness is not high, which indicates that the qualification of enterprises is still essential in innovative elderly care services. The qualification of elderly care enterprises is relatively neglected and needs attention.

In terms of economy, the importance and expressiveness of high-quality employees and guaranteed quality of pension products are relatively high, which indicates that in the intelligent supply chain of Chinese innovative pension services, the quality of pension employees and products is currently in line with the status quo of pension in China, and the results are satisfactory so that it can be maintained. The importance and expressiveness of the service profit guarantee of the elderly care industry are relatively low. This paper believes that, at present, the intelligent elderly care service industry in China is in the initial stage of development, and the service profit obtained by it is at a low stage. It is still necessary to guarantee the most basic elderly care service profit.

In terms of society, the large base of elderly care subjects and the high importance and expressiveness of the social elderly care industry indicate that the current China elderly care service intelligent supply chain system is still perfect for such a large number of elderly care groups, and the number of social elderly care enterprises is also increasing. To ease the burden of elderly care, it needs to continue to maintain. The rapid development of China smart elderly care is of low importance but high expressiveness, which indicates that the development of the China innovative elderly care service supply chain is in a relatively rapid stage of development. The importance of the acceptance of target objects is relatively high, but the expressiveness is not high, which indicates that the acceptance of target objects to intelligent pensions is not high. China should increase the publicity of intelligent pensions and the acceptance of target objects to smart pensions.

In terms of technology, the importance and expressiveness of the rapid response of cloud computing are relatively high, which indicates that the cloud computing of Chinese intelligent elderly care service supply chain is relatively fast and can perfectly solve the analysis of various data of China competent elderly care. The deepening of artificial intelligence, the wide coverage of mobile communication, and the large scale of extensive data collection are unimportant. However, the expressiveness is very high, which indicates that the three aspects of the China elderly care service intelligent supply chain system perform well and can well solve the current elderly care pressure in China. The importance of the popularization of the Internet of Things is very high. However, its expressiveness is not high, which indicates that the Internet of Things in the intelligent supply chain system of China elderly care services is not very popular, and the popularization of the Internet of Things needs to be strengthened to facilitate the life of the elderly care groups in China.

## 5 Conclusion and Suggestions

### 5.1 Research Conclusion and Suggestions

With the continuously deepening of population aging, the pressure of social elderly care is also increasing year by year, and the issue of elderly care has become a problem that needs to be solved in China and even China. This article analyzes and summarizes the four aspects of building a smart supply chain system for elderly care services in China and identifies the advantages and disadvantages of these four aspects to optimize Chinese smart elderly care service system in the future.

After a literature review and investigation, this study ultimately established four directions, namely from the policy aspect, from the economic aspect, from the social aspect, and the technical aspect. Collect data based on research on supply chain managers, staff serving the elderly care industry, and target audience. Moreover, through reliability and validity analysis, the results show that all four have good overall fitness. Through IPA analysis, the development trends of each aspect are obtained, and the main conclusion is to enhance the target audience’s acceptance of intelligent elderly care. Only in this way can the China elderly care service intelligent supply chain system develop more healthily. At the same time, it is necessary to strengthen the qualification review of elderly care enterprises to prevent damaging enterprises from affecting the intelligent supply chain system of elderly care services in China. For the Internet of Things, it is also necessary to make it more universal to ensure real-time supervision of older people’s health, life, and well-being.

### 5.2 Research inspiration

At present, the intelligent supply chain system for elderly care services in China is in the initial stage of development and has excellent potential for development in the future. In recent years, the country has attached increasing importance to elderly care, which is not only a problem in China, but also a problem that needs to be solved nationwide and globally. By building an intelligent supply chain system for elderly care services in China, it can effectively alleviate the pressure on elderly care, better allocate resources, and rationalize their application. At the same time, this is also highly beneficial for the elderly population, making their lives more convenient and allowing them to feel more happiness.

### 5.3 Research Limitations and future recommendations

Although this study has achieved some results, there are still certain research limitations. Firstly, in terms of methodology, this study only used IPA analysis as an analytical method and did not introduce other analytical methods, which may also lead to slight errors in indicator analysis. Secondly, this study is only based on a small portion of research data, which has significant limitations for China Province. Therefore, this paper suggests that future research should combine various research methods to explore the importance and expressiveness of indicators to obtain more accurate data. At the same time, statistical analysis should be conducted on the data from China to enhance its persuasiveness.

## Data Availability

The data supporting reported results cannot be found.

## Supplementary Materials

There are no available online materials.

## Author Contributions

Conceptualization, You-Yu Dai; methodology, You-Yu Dai; validation, You-Yu Dai; formal analysis, You-Yu Dai; investigation, Guanlong Liu; resources, You-Yu Dai; data curation, You-Yu Dai; writing—original draft preparation, You-Yu Dai and Guanlong Liu; writing—review and editing, You-Yu Dai. All authors have read and agreed to the published version of the manuscript.

## Data Availability Statement

The data supporting reported results cannot be found.

